# Gaps in Sexually Transmitted Infection Screening among Men who Have Sex with Men in PrEP Care in the United States

**DOI:** 10.1101/2020.03.06.20032318

**Authors:** Christina Chandra, Kevin M. Weiss, Colleen F. Kelley, Julia L. Marcus, Samuel M. Jenness

## Abstract

**Background:** The U.S. Centers for Disease Control and Prevention (CDC) recommends comprehensive sexually transmitted infection (STI) screening every 3–6 months for men who have sex with men (MSM) using HIV preexposure prophylaxis (PrEP). The gaps between these recommendations and clinical practice by region have not been quantified.

**Methods:** We used survey data collected from the internet-based ARTnet study between 2017 and 2019 on STI screening among MSM across the U.S., stratified by current, prior, and never PrEP use. Poisson regression models with robust error variance were used to model factors, including residence in the Southeast, associated with consistent (“always” or “sometimes”) exposure site-specific STI screening during PrEP care.

**Results:** Of 3259 HIV-negative MSM, 19% were currently using PrEP, 6% had used PrEP in the past, and 75% had never used PrEP. Among ever PrEP users, 87%, 78%, 57%, and 64% reported consistent screening for STIs by blood sample, urine sample or urethral swab, rectal swab, or pharyngeal swab, respectively, during PrEP care. Compared to PrEP users in all other regions, PrEP users in the Southeast were significantly less likely to be consistently screened for urogenital (adjusted prevalence ratio [aPR], 0.86; 95% confidence interval [CI], 0.76–0.98) and rectal STIs (aPR, 0.76; 95% CI, 0.62–0.93) during PrEP care.

**Conclusions:** Substantial gaps exist between CDC recommendations for STI screening during PrEP care and current clinical practice, particularly for rectal and pharyngeal exposure sites that can harbor asymptomatic infections and for MSM in Southeast states where the STI burden is substantial.

**SUMMARY:** Nearly half of U.S. men who have sex with men in PrEP care are not receiving consistent bacterial STI screening at sites of sexual exposure, and levels are worse in the Southeast region where the burden of STI is highest.

## INTRODUCTION

In the United States (U.S.), cases of the major reportable bacterial sexually transmitted infections (STIs) — syphilis, gonorrhea, and chlamydia — increased by up to 71% between 2014 and 2018, reaching all-time highs [1]. Rates of new HIV diagnoses have remained stable overall but increased in key subgroups during this time [2]. Men who have sex with men (MSM) are disproportionately affected by STIs and HIV, accounting for more than half of new infectious syphilis cases, 40% of gonorrhea cases, and 66% of HIV cases in 2018 despite only representing 4% of the population [1–3]. Additionally, more than half of new HIV diagnoses in 2018 occurred in the South, which also has the highest STI rates [1,2].

One hypothesis for the increase in STI cases among MSM has been the growing use of HIV preexposure prophylaxis (PrEP). In a meta-analysis, the pooled STI prevalence among PrEP users within three months of PrEP initiation was 24%, and STI incidence was estimated at 72 per 100 person-years among persistent PrEP users globally [4]. U.S. studies have found that STI prevalence or incidence increased among MSM after PrEP initiation [5,6]. The difference in STI rates before and after PrEP initiation could result from multiple factors, including increased detection of cases from more frequent screening in PrEP care, changes in sexual risk behavior after PrEP initiation (“risk compensation”), and trends in sexual behavior and STI rates that pre-date PrEP [1,7–10]. Many PrEP users are at elevated risk of STIs because PrEP is recommended for MSM with a history of inconsistent condom use and multiple partners [11].

The U.S. Centers for Disease Control and Prevention (CDC) recommends comprehensive STI screening every 3–6 months for MSM using PrEP [11]. This includes blood tests to screen for syphilis, and urine samples and urethral, pharyngeal, and rectal swabs for gonorrhea and chlamydia infections. Prior modeling research suggested that while STI diagnoses may initially increase with expanded PrEP use due to variations of risk compensation, routine STI screening and treatment during PrEP care could prevent over 40% of gonorrhea and chlamydia cases among MSM over ten years [12]. These results were partially attributed to increased treatment of asymptomatic STIs that would have otherwise been undiagnosed. These projections assumed complete adherence to PrEP-related STI screening and treatment recommendations; secondary model scenarios projected that STI incidence could increase if fewer than 50% of PrEP users were consistently screened and treated for STIs.

Quantifying any gaps between PrEP STI screening recommendations and clinical practice is therefore critical to understanding the role of PrEP care in STI control. Several studies have evaluated this in different regions, finding lower rates of STI screening than recommended [7,13–16]. While these studies provide important evidence regarding gaps in PrEP care, there are still no national-level studies of MSM that have compared these gaps across geographic regions. Geographic heterogeneity in clinical practice is important to understand because of the regional differences in infrastructure for PrEP services (e.g., the ability to assess kidney function and serological confirmation of Hepatitis B status for PrEP eligibility) and the spatial clustering of STIs in the Southeast [17,18]. Differences in STI screening practices may also reflect state-level factors that impact PrEP care, including access to healthcare (e.g., Medicaid expansion) and social determinants of health that may vary by state (e.g., housing stability, employment) [19,20]. By examining these differences, we may better understand the role that geographically-focused efforts — such as the federal Ending the HIV Epidemic initiative — may play in stemming both the HIV and STI epidemics in the Southeast [21].

In this study, we compared STI screening among MSM by PrEP use history in a study of MSM in the U.S. We then evaluated factors associated with consistent STI screening at PrEP care visits, including residence in the Southeast and recent sexual exposure at anatomical sites. We hypothesized that there would be strong geographic differences in STI screening consistency among PrEP users even after controlling for demographic and behavioral factors.

## METHODS

### Study Design

ARTnet was a cross-sectional web-based study of MSM in the U.S. conducted between 2017 and 2019 [22]. The study recruited participants through the national American Men’s Internet Survey (AMIS), with a response rate of 53% (4904/9295) [23]. ARTnet eligibility criteria included male sex at birth, current male identity, lifetime history of sexual activity with another man, and age between 15 and 65 years. The study, provided only in English, surveyed participants about recent sexual behaviors, HIV and STI screening, and egocentric network data [24]. We restricted the participants in this analysis to PrEP-eligible MSM, defined as MSM who had ever had an HIV test and who self-reported as HIV-negative. The Emory University Institutional Review Board reviewed and approved the study.

### Measures

We quantified participant-reported data on HIV and STI screening from blood samples, urine sample/urethral swabs, and rectal and pharyngeal swabs at PrEP care visits and any HIV or STI screening in the past 12 months. Participants who had ever used PrEP were asked: “At these PrEP (i.e., Truvada) check-up visits, how often did you get tested for HIV and sexually transmitted diseases (STD)? STDs could include gonorrhea, chlamydia, and syphilis.” Participants could answer “Always”, “Sometimes”, “Rarely”, or “Never” for HIV screening or STI screening site/method: throat (pharyngeal swab), anus (rectal swab), genital/penis (urine sample/urethral swab), or blood sample. We assumed blood-based STI screening was used to possibly screen for syphilis, since blood samples are necessary for syphilis screening while swabs and urine samples are used for nucleic acid amplification tests (NAATs), the gold standard for gonorrhea and chlamydia testing. For statistical models, we dichotomized the outcomes to consistent screening (“Always” or “Sometimes”) versus inconsistent screening (“Rarely” or “Never”).

We then investigated correlates of consistent STI screening. We considered age group, race/ethnicity, current health insurance, annual household income, recent sexual exposure at anatomical site, and geographic region of residence. Using data on the five most recent reported partners in the past year, respondents were classified as having been exposed at the urethra (any insertive anal or insertive oral intercourse), rectum (any receptive anal intercourse), or pharynx (any receptive oral intercourse). For geography, we considered census division and state by matching reported residential ZIP Codes against census data [22]. Our primary exposure of interest was residence in 12 Southeast states as defined by the U.S. Bureau of Economic Analysis: Alabama, Arkansas, Florida, Georgia, Kentucky, Louisiana, Mississippi, North Carolina, South Carolina, Tennessee, Virginia, and West Virginia [25].

### Statistical Analysis

Our primary research objective was to quantify the association between residence in Southeast states and consistent screening for STIs among MSM who were currently using or had previously used PrEP. We first estimated the bivariable prevalence ratios between our hypothesized correlates — age, race/ethnicity, current health insurance, annual household income, recent sexual exposure at anatomical site, and geography — and consistent site-specific STI screening using Poisson regression with robust error variance and complete case analysis. In multivariable Poisson regression models, we estimated the prevalence ratios between our primary exposure (residence in a Southeast state) and consistent STI screening adjusted for four variables that we hypothesized would confound this relationship: age, race/ethnicity, annual household income, and sexual exposure at anatomical site. We did not control for variables that we considered mediators of the associations, such as health insurance that may vary by state as a result of Medicaid expansion. All analyses were conducted in R 3.6.1 [26]. Analysis scripts are provided in a GitHub repository (https://github.com/EpiModel/PrEP-STI-Test), and the primary data are available upon request.

## RESULTS

After excluding PrEP-ineligible participants, 3259 MSM were included in this analysis. Of those, 631 (19%) were currently using PrEP at the time of survey completion, 178 (6%) had previously used PrEP, and 2450 (75%) had never used PrEP. **Table 1** presents the demographics and HIV/STI screening history by PrEP use. Overall, 74% were non-Hispanic white, 51% were aged 15-34 years, and 23% resided in Southeast states. Residents in the Southeast were generally underrepresented, accounting for 25% of MSM who had never used PrEP and 19% of both current and prior PrEP users. Four-fifths (82%) of current PrEP users reported having any private health insurance compared with 72% of prior PrEP users and 75% of never PrEP users. Only 3% of current PrEP users were uninsured compared with 10% of prior PrEP users and 9% of never PrEP users. A higher proportion of current PrEP users reported screening for a bacterial STI (without specifying specimen collected) in the past year (89%), compared to prior PrEP users (71%) and never PrEP users (44%). Similarly, 98% of current PrEP users were screened for HIV in the past year compared to 80% of prior PrEP users and 65% of never PrEP users.

**Table 1.** Characteristics of PrEP-Eligible ARTnet Participants, Stratified by PrEP Use History

|  | **Total<sup>a</sup>** |  | **Current PrEP Use** |  | **Prior PrEP Use** |  | **Never Used PrEP** |  |
| --- | --- | --- | --- | --- | --- | --- | --- | --- |
|  | N or Mean | %<sup>b</sup> or SD | N or Mean | % or SD | N or Mean | % or SD | N or Mean | % or SD |
| **Total Sample** | 3259 | 100.0 | 631 | 100.0 | 178 | 100.0 | 2450 | 100.0 |
| **Race/Ethnicity** |  |  |  |  |  |  |  |  |
| Black (Non-Hispanic) | 134 | 4.1 | 28 | 4.4 | 4 | 2.3 | 102 | 4.2 |
| Hispanic | 422 | 12.9 | 81 | 12.8 | 23 | 12.9 | 318 | 13.0 |
| Other (Non-Hispanic) | 283 | 8.7 | 53 | 8.4 | 11 | 6.2 | 219 | 8.9 |
| White (Non-Hispanic) | 2420 | 74.3 | 469 | 74.3 | 140 | 78.7 | 1811 | 73.9 |
| **Age** | 37.3 | 13.6 | 39.5 | 12.4 | 34.1 | 11.5 | 36.9 | 13.9 |
| 15–24 | 713 | 21.9 | 71 | 11.3 | 36 | 20.2 | 606 | 24.7 |
| 25–34 | 947 | 29.1 | 196 | 31.1 | 76 | 42.7 | 675 | 27.6 |
| 35–44 | 518 | 15.9 | 122 | 19.3 | 26 | 14.6 | 370 | 16.3 |
| 45–54 | 573 | 17.6 | 149 | 23.6 | 24 | 13.5 | 400 | 16.3 |
| 55–65 | 508 | 15.6 | 93 | 14.7 | 16 | 9.0 | 399 | 16.3 |
| **Residence in Census Division<sup>c</sup>** |  |  |  |  |  |  |  |  |
| Pacific | 562 | 17.2 | 155 | 24.6 | 31 | 17.4 | 376 | 15.3 |
| Mountain | 281 | 8.6 | 30 | 4.8 | 21 | 11.8 | 230 | 9.4 |
| West North Central | 187 | 5.7 | 32 | 5.1 | 13 | 7.3 | 142 | 5.8 |
| East North Central | 466 | 14.3 | 92 | 14.6 | 25 | 14.0 | 349 | 14.2 |
| West South Central | 323 | 9.9 | 61 | 9.7 | 11 | 6.2 | 251 | 10.2 |
| East South Central | 132 | 4.1 | 19 | 3.0 | 6 | 3.4 | 107 | 4.4 |
| South Atlantic | 701 | 21.5 | 119 | 18.9 | 35 | 19.7 | 547 | 22.3 |
| Middle Atlantic | 442 | 13.6 | 92 | 14.6 | 20 | 11.2 | 330 | 13.5 |
| New England | 165 | 5.1 | 31 | 4.9 | 16 | 9.0 | 118 | 4.8 |
| **Residence in Southeast<sup>b</sup>** |  |  |  |  |  |  |  |  |
| Yes | 760 | 23.3 | 121 | 19.2 | 33 | 18.5 | 606 | 24.7 |
| No | 2499 | 76.7 | 510 | 80.8 | 145 | 81.5 | 1844 | 75.3 |
| **Health Insurance** |  |  |  |  |  |  |  |  |
| Private | 2441 | 76.4 | 516 | 82.2 | 128 | 71.9 | 1797 | 75.2 |
| Public | 514 | 16.1 | 94 | 15.0 | 33 | 18.5 | 387 | 16.2 |
| None | 242 | 7.6 | 18 | 2.9 | 17 | 9.6 | 207 | 8.7 |
| **Annual Household Income** |  |  |  |  |  |  |  |  |
| \$0 to \$19,999 | 372 | 12.3 | 54 | 9.3 | 18 | 10.7 | 300 | 13.2 |
| \$20,000 to \$39,999 | 574 | 19.1 | 93 | 16.1 | 34 | 20.2 | 447 | 19.7 |
| \$40,000 to \$74,999 | 861 | 28.6 | 153 | 26.4 | 56 | 33.3 | 652 | 28.8 |
| \$75,000 or more | 1206 | 40.0 | 279 | 48.2 | 60 | 35.7 | 867 | 38.3 |
| **Screened for HIV in the Past 12 Months** |  |  |  |  |  |  |  |  |
| Yes | 2106 | 71.9 | 561 | 98.2 | 129 | 79.6 | 1416 | 64.5 |
| No | 822 | 28.1 | 10 | 1.8 | 33 | 20.4 | 779 | 35.5 |
| **Screened for an STI in Past 12 Months** |  |  |  |  |  |  |  |  |
| Yes | 1677 | 54.4 | 547 | 89.1 | 122 | 70.9 | 1008 | 43.8 |
| No | 1408 | 45.6 | 67 | 10.9 | 50 | 28.1 | 1291 | 56.2 |

Abbreviations: PrEP, preexposure prophylaxis; SD, standard deviation; USD, U.S. Dollars; STI, sexually transmitted infection
<sup>a</sup> Totals may not add up to sample total due to missing data for health insurance, annual household income, and screening for
<sup>b</sup> Column percents
<sup>c</sup> https://www2.census.gov/geo/pdfs/maps-data/maps/reference/us_regdiv.pdf
<sup>d</sup> Residence in Alabama, Arkansas, Florida, Georgia, Kentucky, Louisiana, Mississippi, North Carolina, South Carolina, Tennessee, Virginia, and West Virginia

Among MSM who had ever used PrEP, 84% of current users reported attending PrEP care visits at least every three months compared to 71% of prior PrEP users (**Table 2**). Overall, 91% of MSM who had ever used PrEP were always screened for HIV at their PrEP care visits. Greater proportions of current PrEP users, compared to prior PrEP users, reported “always” being screened for STIs by throat swab (44% versus 35%), rectal swab (37% versus 32%), blood (possibly for syphilis; 70% versus 57%), and urine sample/urethral swab (59% versus 50%) at PrEP care visits. In contrast, MSM who had ever used PrEP had also reported “never” providing a rectal swab (35%), throat swab (28%), and urine sample/urethral swab (17%) for STI screening during PrEP care visits.

**Table 2.**
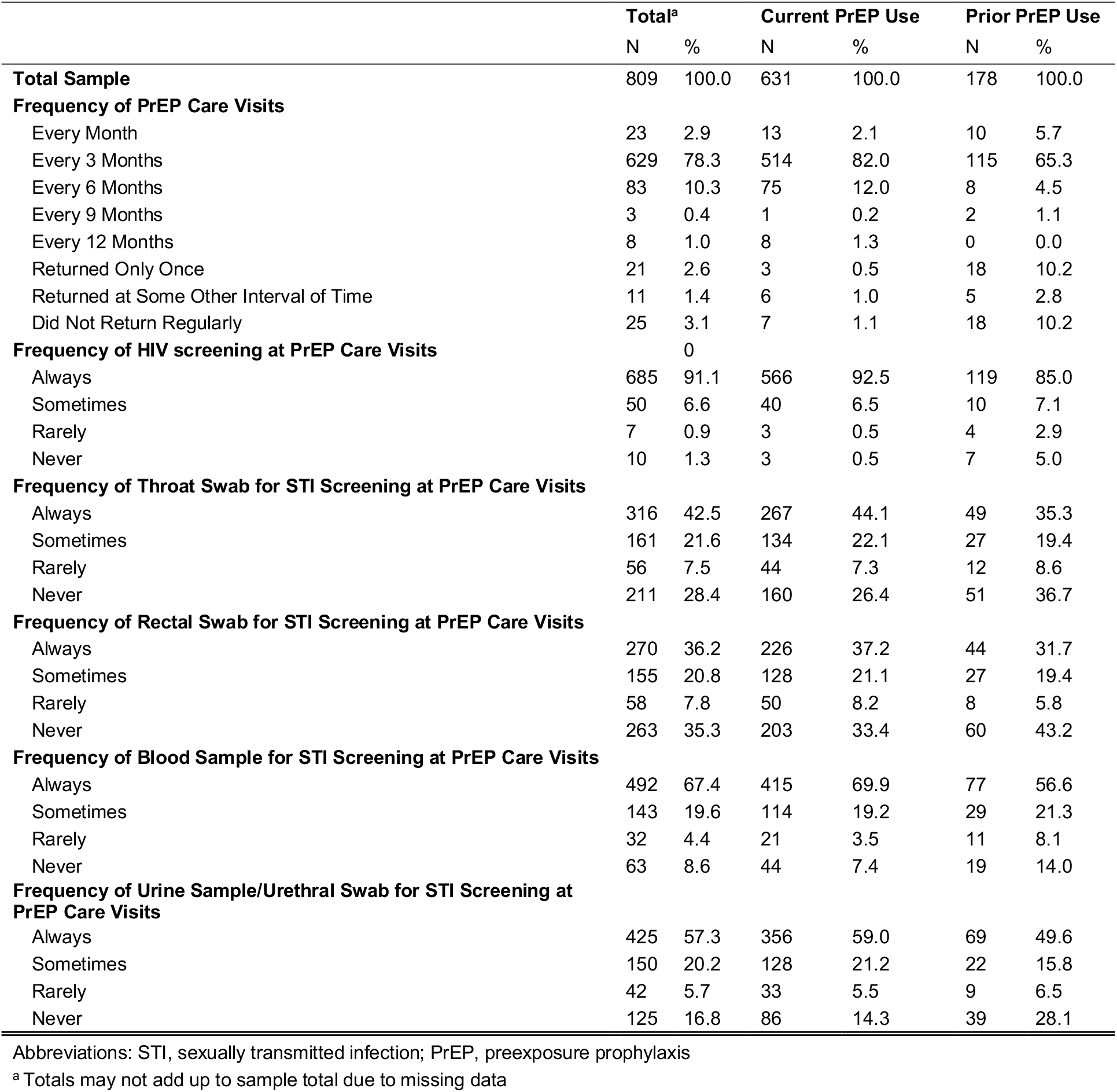
Frequency of PrEP Care Visits and HIV/STI Screening at PrEP Care Visits among MSM with Current or Prior PrEP Use

A higher percentage of MSM who had ever used PrEP reported consistent possible screening for syphilis (87%) and urogenital STI screening (78%) (**Table 3**) compared to pharyngeal (64%) and rectal STI screening (57%) (**Table 4**). Compared to MSM aged 15-24 years, older MSM aged 55-65 years had the lowest levels of consistent screening for urogenital STIs (80% versus 71%), pharyngeal STIs (70% versus 60%), and rectal STIs (67% versus 53%) (**Tables 3 and 4**). Consistent screening was reported less in Southeast states for urogenital STI screening (66% versus 80% in all other states), pharyngeal STI screening (55% versus 66%), and rectal STI screening (44% versus 60%). Respondents reporting recent exposure at corresponding anatomical sites also reported more consistent screening for urogenital (78% versus 68% among those unexposed) and rectal STIs (60% versus 44%). Proportions of MSM reporting consistent screening of pharyngeal STIs were about the same between MSM reporting sexual exposure and none at the pharynx (64% versus 63%).

**Table 3.** Bivariable Associations of Consistent Possible Syphilis and Urogenital STI Screening with Demographic and Behavioral Factors among MSM Who Have Ever Used PrEP

|  | Possible Syphilis Screening<sup>a</sup> (N=730) |  |  |  |  |  | Urogenital STI Screening (N=742) |  |  |  |  |  |
| --- | --- | --- | --- | --- | --- | --- | --- | --- | --- | --- | --- | --- |
|  | Consistent<sup>b</sup> |  | Inconsistent |  | PR | 95% CI | Consistent |  | Inconsistent |  | PR | 95% CI |
|  | N | % | N | % |  |  | N | % | N | % |  |  |
| **Total Sample** | 635 | 87.0 | 95 | 13.0 |  |  | 575 | 77.5 | 167 | 22.5 |  |  |
| **Age** |  |  |  |  |  |  |  |  |  |  |  |  |
| 15-24 | 75 | 83.3 | 15 | 16.7 | 1.00 | - | 73 | 80.2 | 18 | 19.8 | 1.00 | - |
| 25-34 | 215 | 87.8 | 30 | 12.2 | 1.05 | 0.95, 1.17 | 201 | 80.1 | 50 | 19.9 | 1.00 | 0.89, 1.12 |
| 35-44 | 123 | 90.4 | 13 | 9.6 | 1.09 | 0.97, 1.21 | 104 | 75.4 | 34 | 24.6 | 0.94 | 0.82, 1.08 |
| 45-54 | 140 | 85.9 | 23 | 14.1 | 1.03 | 0.92, 1.15 | 127 | 77.9 | 36 | 22.1 | 0.97 | 0.85, 1.11 |
| 55-65 | 82 | 85.4 | 14 | 14.6 | 1.02 | 0.91, 1.16 | 70 | 70.7 | 29 | 29.3 | 0.88 | 0.75, 1.04 |
| **Race/Ethnicity** |  |  |  |  |  |  |  |  |  |  |  |  |
| Black (Non-Hispanic) | 30 | 93.8 | 2 | 6.3 | 1.00 | - | 28 | 87.5 | 4 | 12.5 | 1.00 | - |
| Hispanic | 84 | 91.3 | 8 | 8.7 | 0.97 | 0.87, 1.09 | 78 | 84.8 | 14 | 15.2 | 0.97 | 0.83, 1.13 |
| Other (Non-Hispanic) | 53 | 91.4 | 5 | 8.6 | 0.97 | 0.87, 1.10 | 45 | 77.6 | 13 | 22.4 | 0.89 | 0.73, 1.07 |
| White (Non-Hispanic) | 468 | 85.4 | 80 | 14.6 | 0.91 | 0.83, 1.00 | 424 | 75.7 | 136 | 24.3 | 0.87 | 0.75, 0.99 |
| **Health Insurance** |  |  |  |  |  |  |  |  |  |  |  |  |
| None | 28 | 84.8 | 5 | 15.2 | 1.00 | - | 23 | 69.7 | 10 | 30.3 | 1.00 | - |
| Public | 98 | 87.5 | 14 | 12.5 | 1.03 | 0.88, 1.21 | 91 | 79.8 | 23 | 20.2 | 1.15 | 0.90, 1.46 |
| Private | 507 | 87.1 | 75 | 12.9 | 1.03 | 0.89, 1.19 | 459 | 77.5 | 133 | 22.5 | 1.11 | 0.88, 1.40 |
| **Annual Household Income** |  |  |  |  |  |  |  |  |  |  |  |  |
| \$0 to \$19,999 | 54 | 83.1 | 11 | 16.9 | 1.00 | - | 50 | 76.9 | 15 | 23.1 | 1.00 | - |
| \$20,000 to \$39,999 | 86 | 80.4 | 21 | 19.6 | 0.97 | 0.84, 1.12 | 87 | 79.1 | 23 | 20.9 | 1.03 | 0.87, 1.21 |
| \$40,000 to \$74,999 | 165 | 87.3 | 24 | 12.7 | 1.05 | 0.93, 1.19 | 147 | 75.4 | 48 | 24.6 | 0.98 | 0.84, 1.14 |
| \$75,000 or more | 279 | 89.1 | 34 | 10.9 | 1.07 | 0.96, 1.21 | 244 | 77.2 | 72 | 22.8 | 1.00 | 0.87, 1.16 |
| **Residence in Census Division** |  |  |  |  |  |  |  |  |  |  |  |  |
| New England | 36 | 92.3 | 3 | 7.7 | 1.00 | - | 33 | 80.5 | 8 | 19.5 | 1.00 | - |
| Middle Atlantic | 89 | 85.6 | 15 | 14.4 | 0.93 | 0.82, 1.05 | 86 | 79.6 | 22 | 20.4 | 0.99 | 0.83, 1.18 |
| East North Central | 89 | 84.0 | 17 | 16.0 | 0.91 | 0.80, 1.03 | 83 | 77.6 | 24 | 22.4 | 0.96 | 0.80, 1.16 |
| West North Central | 29 | 74.4 | 10 | 25.6 | 0.81 | 0.66, 0.99 | 24 | 60.0 | 16 | 40.0 | 0.75 | 0.56, 1.00 |
| South Atlantic | 119 | 86.9 | 18 | 13.1 | 0.94 | 0.84, 1.05 | 93 | 67.9 | 44 | 32.1 | 0.84 | 0.70, 1.02 |
| East South Central | 18 | 85.7 | 3 | 14.3 | 0.93 | 0.76, 1.13 | 13 | 61.9 | 8 | 38.1 | 0.77 | 0.53, 1.11 |
| West South Central | 62 | 89.9 | 7 | 10.1 | 0.97 | 0.86, 1.10 | 57 | 82.6 | 12 | 17.4 | 1.03 | 0.85, 1.24 |
| Mountain | 34 | 79.1 | 9 | 20.9 | 0.86 | 0.72, 1.02 | 32 | 74.4 | 11 | 25.6 | 0.92 | 0.73, 1.17 |
| Pacific | 159 | 92.4 | 13 | 7.6 | 1.00 | 0.91, 1.11 | 154 | 87.5 | 22 | 12.5 | 1.09 | 0.93, 1.28 |
| **Residence in Southeast<sup>c</sup>** |  |  |  |  |  |  |  |  |  |  |  |  |
| No | 517 | 87.2 | 76 | 12.8 | 1.00 | - | 484 | 80.0 | 121 | 20.0 | 1.00 | - |
| Yes | 118 | 86.1 | 19 | 13.9 | 0.99 | 0.92, 1.06 | 91 | 66.4 | 46 | 33.6 | 0.83 | 0.73, 0.94 |
| **Exposure at Anatomical Site<sup>d</sup>** |  |  |  |  |  |  |  |  |  |  |  |  |
| No |  |  |  |  |  |  | 15 | 68.2 | 7 | 31.8 | 1.00 | - |
| Yes |  |  |  |  |  |  | 560 | 77.8 | 160 | 22.2 | 1.14 | 0.86, 1.52 |

Abbreviations: STI, sexually transmitted infection; PrEP, preexposure prophylaxis; PR, prevalence ratio; CI, confidence interval
<sup>a</sup> Participants who provided blood samples for STI screening were considered to be possibly screened for syphilis, since blood samples are needed for syphilis screening, while swabs and urine samples can be used for gold standard testing of gonorrhea and chlamydia
<sup>b</sup> Consistent screening defined as always or sometimes (versus rarely or never) being screened at PrEP care visits compared
<sup>c</sup> Alabama, Arkansas, Florida, Georgia, Kentucky, Louisiana, Mississippi, North Carolina, South Carolina, Tennessee, Virginia, and W Virginia
<sup>d</sup> Defined as any insertive anal intercourse or receiving oral sex for urogenital STI screening

**Table 4.** Bivariable Associations of Consistent Rectal and Pharyngeal STI Screening with Demographic and Behavioral Factors among MSM Who Have Ever Used PrEP

|  | Rectal STI Screening (N=746) |  |  |  |  |  | Pharyngeal STI Screening (N=744) |  |  |  |  |  |
| --- | --- | --- | --- | --- | --- | --- | --- | --- | --- | --- | --- | --- |
|  | Consistent |  | Inconsistent |  | PR | 95% CI | Consistent |  | Inconsistent |  | PR | 95% CI |
|  | N | % | N | % |  |  | N | % | N | % |  |  |
| **Total Sample** | 425 | 57.0 | 321 | 43.0 |  |  | 477 | 64.1 | 267 | 35.9 |  |  |
| **Age** |  |  |  |  |  |  |  |  |  |  |  |  |
| 15-24 | 61 | 67.0 | 30 | 33.0 | 1.00 | - | 63 | 70.0 | 27 | 30.0 | 1.00 | - |
| 25-34 | 146 | 57.9 | 106 | 42.1 | 0.86 | 0.72, 1.03 | 163 | 64.9 | 88 | 35.1 | 0.93 | 0.79 1.09 |
| 35-44 | 74 | 53.6 | 64 | 46.4 | 0.80 | 0.65, 0.99 | 89 | 64.5 | 49 | 35.5 | 0.92 | 0.77 1.11 |
| 45-54 | 91 | 55.2 | 74 | 44.8 | 0.82 | 0.67, 1.00 | 102 | 61.8 | 63 | 38.2 | 0.88 | 0.74 1.06 |
| 55-65 | 53 | 53.0 | 47 | 47.0 | 0.79 | 0.63, 1.00 | 60 | 60.0 | 40 | 40.0 | 0.86 | 0.70 1.06 |
| **Race/Ethnicity** |  |  |  |  |  |  |  |  |  |  |  |  |
| Black (Non-Hispanic) | 16 | 50.0 | 16 | 50.0 | 1.00 | - | 19 | 59.4 | 13 | 40.6 | 1.00 | - |
| Hispanic | 57 | 62.0 | 35 | 38.0 | 1.24 | 0.85, 1.82 | 61 | 66.3 | 31 | 33.7 | 1.12 | 0.81, 1.54 |
| Other (Non-Hispanic) | 39 | 67.2 | 19 | 32.8 | 1.34 | 0.91, 1.99 | 43 | 74.1 | 15 | 25.9 | 1.25 | 0.90, 1.73 |
| White (Non-Hispanic) | 313 | 55.5 | 251 | 44.5 | 1.11 | 0.78, 1.58 | 354 | 63.0 | 208 | 37.0 | 1.06 | 0.79, 1.42 |
| **Health Insurance<sup>a</sup>** |  |  |  |  |  |  |  |  |  |  |  |  |
| None | 19 | 57.6 | 14 | 42.4 | 1.00 | - | 22 | 66.7 | 11 | 33.3 | 1.00 | - |
| Public | 58 | 50.0 | 58 | 50.0 | 0.87 | 0.62, 1.23 | 74 | 63.8 | 42 | 36.2 | 0.96 | 0.73, 1.26 |
| Private | 347 | 58.4 | 247 | 41.6 | 1.01 | 0.75, 1.37 | 380 | 64.2 | 212 | 35.8 | 0.96 | 0.75, 1.23 |
| **Annual Household Income** |  |  |  |  |  |  |  |  |  |  |  |  |
| \$0 to \$19,999 | 35 | 53.0 | 31 | 47.0 | 1.00 | - | 44 | 66.7 | 22 | 33.3 | 1.00 | - |
| \$20,000 to \$39,999 | 59 | 53.6 | 51 | 46.4 | 1.01 | 0.76, 1.35 | 62 | 56.4 | 48 | 43.6 | 0.85 | 0.67, 1.07 |
| \$40,000 to \$74,999 | 109 | 55.9 | 86 | 44.1 | 1.05 | 0.81, 1.37 | 125 | 64.4 | 69 | 35.6 | 0.97 | 0.79, 1.18 |
| \$75,000 or more | 187 | 58.8 | 131 | 41.2 | 1.11 | 0.87, 1.42 | 206 | 65.0 | 111 | 35.0 | 0.97 | 0.81, 1.18 |
| **Residence in Census Division** |  |  |  |  |  |  |  |  |  |  |  |  |
| New England | 29 | 70.7 | 12 | 29.3 | 1.00 | - | 30 | 73.2 | 11 | 26.8 | 1.00 | - |
| Middle Atlantic | 65 | 60.2 | 43 | 39.8 | 0.85 | 0.66, 1.09 | 74 | 69.2 | 33 | 30.8 | 0.95 | 0.76, 1.18 |
| East North Central | 57 | 52.8 | 51 | 47.2 | 0.75 | 0.57, 0.97 | 59 | 54.6 | 49 | 45.4 | 0.75 | 0.58, 0.96 |
| West North Central | 21 | 52.5 | 19 | 47.5 | 0.74 | 0.52, 1.06 | 24 | 61.5 | 15 | 38.5 | 0.84 | 0.62, 1.15 |
| South Atlantic | 65 | 46.1 | 76 | 53.9 | 0.65 | 0.50, 0.85 | 79 | 56.0 | 62 | 44.0 | 0.77 | 0.60, 0.97 |
| East South Central | 7 | 33.3 | 14 | 66.7 | 0.47 | 0.25, 0.89 | 9 | 42.9 | 12 | 57.1 | 0.59 | 0.35, 0.99 |
| West South Central | 40 | 58.0 | 29 | 42.0 | 0.82 | 0.62, 1.09 | 47 | 68.1 | 22 | 31.9 | 0.93 | 0.73, 1.19 |
| Mountain | 19 | 45.2 | 23 | 54.8 | 0.64 | 0.43, 0.94 | 20 | 47.6 | 22 | 52.4 | 0.65 | 0.45, 0.94 |
| Pacific | 122 | 69.3 | 54 | 30.7 | 0.98 | 0.79, 1.22 | 135 | 76.7 | 41 | 23.3 | 1.05 | 0.86, 1.28 |
| **Residence in Southeast<sup>b</sup>** |  |  |  |  |  |  |  |  |  |  |  |  |
| No | 364 | 60.1 | 242 | 39.9 | 1.00 | - | 400 | 66.2 | 204 | 33.8 | 1.00 | - |
| Yes | 61 | 43.6 | 79 | 56.4 | 0.73 | 0.59, 0.89 | 77 | 55.0 | 63 | 45.0 | 0.83 | 0.71, 0.97 |
| **Exposure at Anatomical Site<sup>c</sup>** |  |  |  |  |  |  |  |  |  |  |  |  |
| No | 61 | 44.2 | 77 | 55.8 | 1.00 | - | 15 | 62.5 | 9 | 37.5 | 1.00 | - |
| Yes | 364 | 59.9 | 244 | 40.1 | 1.35 | 1.11, 1.65 | 462 | 64.2 | 258 | 35.8 | 1.03 | 0.75, 1.41 |

Abbreviations: STI, sexually transmitted infection; PrEP, preexposure prophylaxis; PR, prevalence ratio; CI, confidence interval
<sup>a</sup> Consistent screening defined as always or sometimes (versus rarely or never) being screened at PrEP care visits compared<sup>b</sup> Alabama, Arkansas, Florida, Georgia, Kentucky, Louisiana, Mississippi, North Carolina, South Carolina, Tennessee, Virginia, and W Virginia<sup>c</sup> Defined as any receptive anal intercourse for rectal STI screening and any performance of oral sex on a partner for pharyngeal STI screening within the past year

Among MSM who had ever used PrEP, age, race/ethnicity, health insurance, annual household income, census division, and residence in Southeast states were not significantly associated with possible syphilis screening at PrEP care visits (**Table 3**). For urogenital, rectal, and pharyngeal STIs, there was a general trend of MSM aged 15-24 years reporting consistent screening compared to MSM aged 55-65 years at PrEP care visits (**Tables 3 and 4**). In unadjusted analyses, MSM in Southeast states reported a lower prevalence of consistent urogenital (prevalence ratio [PR], 0.83; 95% confidence interval [CI], 0.73–0.94), rectal (PR, 0.73; 95% CI, 0.59–0.89), and pharyngeal (PR, 0.83; 95% CI, 0.71–0.97) STI screening during PrEP care (**Table 4**). MSM who reported exposure at the rectum reported a 35% higher prevalence of consistent rectal STI screening at PrEP care visits (PR, 1.35; 95% CI, 1.11–1.65). Prevalence of consistent STI screening was not substantially different between MSM reporting exposure at anatomical site compared to those with no exposure for urogenital (PR, 1.14; 95% CI, 0.86–1.52) and pharyngeal STIs (PR, 1.03; 95% CI, 0.75–1.41) (**Tables 3 and 4**).

In adjusted models, a smaller proportion of MSM in Southeast states reported consistent STI screening compared to MSM in other regions (**Table 5**). MSM in the Southeast had 14% lower prevalence of consistent urogenital STI screening (adjusted prevalence ratio [aPR], 0.86; 95% CI, 0.76– 0.98) and 24% lower prevalence of consistent rectal STI screening (aPR, 0.76; 95% CI, 0.62–0.93) during PrEP care. A lower prevalence of MSM in Southeast states compared to all other regions of the country reported consistent pharyngeal STI screening during PrEP care (aPR, 0.87; 95% CI, 0.74–1.03). The prevalence of consistent possible syphilis screening was not substantially different between MSM in the Southeast and MSM in all other regions (aPR, 1.02; 95% CI, 0.95–1.10).

**Table 5.** Multivariable Associations of Consistent^a^ STI Screening with Residence in the Southeast U.S.^b^ Among MSM who Have Ever Used PrEP

|  | Adjusted PR<sup>c</sup> | 95% CI |
| --- | --- | --- |
| **Possible Syphilis Screening<sup>d,e</sup>** |  |  |
| Residence Outside Southeast | 1.00 | - |
| Residence in Southeast | 1.02 | 0.95, 1.10 |
| **Urogenital STI Screening** |  |  |
| Residence Outside Southeast | 1.00 | - |
| Residence in Southeast | 0.86 | 0.76, 0.98 |
| **Rectal STI Screening** |  |  |
| Residence Outside Southeast | 1.00 | - |
| Residence in Southeast | 0.76 | 0.62, 0.93 |
| **Pharyngeal STI Screening** |  |  |
| Residence Outside Southeast | 1.00 | - |
| Residence in Southeast | 0.87 | 0.74, 1.03 |

Abbreviations: STI, sexually transmitted infection; PrEP, preexposure prophylaxis; PR, prevalence ratio; CI, confidence interval
<sup>a</sup> Consistent screening defined as always or sometimes (versus rarely or never) being screened at PrEP care visits compared
<sup>b</sup> Residence in Alabama, Arkansas, Florida, Georgia, Kentucky, Louisiana, Mississippi, North Carolina, South Carolina, Tennessee, Virginia, and West Virginia
<sup>c</sup> Adjusted for age, race/ethnicity, annual household income, and sexual exposure at anatomical site
<sup>d</sup> Not adjusted for sexual exposure at anatomical site
<sup>e</sup> Participants who provided blood samples for STI screening were considered to be possibly screened for syphilis, since blood samples are needed for syphilis screening, while swabs and urine samples can be used for gold standard testing of gonorrhea and chlamydia

## DISCUSSION

In this national study of MSM in the U.S., consistency of STI screening at PrEP care visits was lower than recommended, especially for rectal and pharyngeal infections that are mostly asymptomatic. Our findings also highlight the regional variation in gaps between recommendations and PrEP clinical practice overall, and raise concerns about whether comprehensive PrEP care as currently practiced would be effective for STI control, particularly in the Southeast where the burden of bacterial STIs is highest [1,18].

This study contributes to the growing literature on patterns of STI screening within PrEP care. With the exception of one national study [14], research has been limited to individual clinics or health systems, often with limited sample sizes [7,13,15,16]. These studies analyze a variety of data streams, including electronic health records, health insurance claims, or laboratory screening [15,16,27]. Many different measures have been used as primary outcomes: screening at last PrEP care visit [13]; STI or rectal STI screening in the prior 12 months [7]; or proportion screened for STIs within 6 months of PrEP initiation [14]. Our study, in contrast, measured STI screening *across* PrEP care visits that may better reflect ongoing consistency of clinical practice. We also stratified data by current and prior PrEP use, and more frequent STI screening among current PrEP users compared to prior users was expected given CDC’s revised recommendations to more frequent STI screening in 2017.

Despite these different study designs and outcomes, research has consistently shown suboptimal STI screening among PrEP users, especially for extragenital (rectal or pharyngeal) screening [13–16]. Similar to a study conducted in Baltimore, we found that among all site-specific screening, rectal swabs were reported least often [16]. A study in New York City, in contrast, found that pharyngeal STI screening was the least reported (at the last PrEP care visit) [13]. These results are also consistent with findings of infrequent extragenital STI screening among MSM *outside* of PrEP care [28–30]. Infrequent extragenital screening is a public health concern because of the higher prevalence of asymptomatic chlamydia or gonorrhea infections that may remain undetected [31]. Asymptomatic STIs at these sites also lead to increased risk of HIV infection among partners in the sexual network not using PrEP [24,32].

Several factors have been found to be associated with more recent or comprehensive STI screening: younger age, white race, college education, reporting exposure at anatomical sites that may trigger screening, and previous syphilis diagnoses [7,13,15,16]. We additionally found that residence in the Southeast was associated with inconsistent urogenital and rectal STI screening, after adjusting for potential confounders. These results have implications for regional improvements of comprehensive PrEP care at the patient, provider, and systems level. MSM have previously noted discomfort in discussing their sexual orientation and subsequently sexual health and behaviors with their primary care providers, emphasizing the importance of patient-provider communication in PrEP care [33]. At the provider level, time constraints, cultural and language barriers, difficulty obtaining a sexual history, and patient privacy concerns have affected HIV providers’ ability to conduct routine STI screening [34]. The cost of ancillary services associated with PrEP care such as STI screening may also be a barrier, as providers have noted that high cost of medication and health insurance coverage may hinder the real-world effectiveness of PrEP [35]. Related to cost, alternative options should also be considered for undocumented populations. Moreover, 7 of the 12 Southeast states (in our definition) have not expanded Medicaid under the Affordable Care Act, potentially limiting the ability of MSM to obtain adequate coverage for consistent STI screening [36]. Gaps in comprehensive PrEP care may exacerbate the large racial and socioeconomic disparities in HIV and STI incidence in the region [37]. Wider implementation of alternative STI screening approaches, such as self-collected rectal and pharyngeal swabs or home-screening kits, may also improve STI screening coverage for PrEP-using MSM [38,39].

### Limitations

This study has several limitations. First, this web-based sample does not necessarily represent the larger population of MSM with respect to age, race/ethnicity, health insurance, annual household income, and geographic residential location. Most participants were white, had higher income levels, had private health insurance, and lived in the mid-Atlantic/Northeast or the West Coast. Compared to venue-based sampling, web-based sampling of MSM may yield a sample with more non-Hispanic white and higher-income men [40]. The survey was English-only, which may have contributed to the lack of ethnic representativeness of the responses. Additionally, given our focus on the Southeast and that 4% of respondents are Black, our sample critically underrepresents the MSM population in the Southeast. Second, stratifying participant data by the covariates of interest yielded small sample sizes in some categories (reflected in wide confidence intervals for some estimates). Third, we relied on self-reported data, and there may be misclassification of screening frequency due to not understanding what specific STIs are being screened. We may be overestimating consistency of possible syphilis screening, because participants may misinterpret blood draws for HIV screening or other tests at PrEP care visits as inclusive of STI screening. Fourth, the data were also limited in classifying anatomical site exposure since the ARTnet study collected data on a maximum of five partners; this may not align with their PrEP care visits. Fifth, we assumed that PrEP users accessed PrEP in their reported residential ZIP Code, but it is possible that prior or current PrEP users accessed PrEP in a different state. Sixth, respondents may have received STI screening outside of PrEP care (e.g., through another healthcare provider or community-based organization), the frequency of which is not captured in ARTnet. Finally, the STI screening frequency measure (always, sometimes, rarely, or never) allows for an approximate but not perfect estimate of adherence to CDC recommendations, particularly because “sometimes” can represent a wide range of screening frequencies as interpreted by respondents. For this reason, we used the term “consistent” rather than “as recommended.”

## Conclusions

Data from this large national study of MSM improved the understanding of geographic differences in comprehensive health services for MSM using PrEP. Although a higher proportion of PrEP users reported recent STI screening compared to non-PrEP users, barriers exist to complete adherence to CDC PrEP guidelines of comprehensive screening for STIs at all exposure sites among MSM. This was especially true in Southeast states where the STI burden is high and where the federal government has prioritized funding HIV prevention programs through the Ending the HIV Epidemic initiative [21]. As PrEP is scaled up in the Southeast through this national initiative, resources at the patient, provider, and systems level to support STI screening and treatment as part of PrEP care should simultaneously be strengthened. Otherwise, suboptimal STI screening in PrEP care may lead to increases in overall STI incidence among MSM.

## Data Availability

Analysis scripts are provided in a Github repository are provided in a repository linked below. Primary data is available for analysis with a data use agreement available from the corresponding author.

https://github.com/EpiModel/PrEP-STI-Test

